# Genetic background shapes skin cancer risk in patients with autoimmune hypothyroidism

**DOI:** 10.64898/2026.10.15.26365654

**Authors:** Ivan Molotkov, Mary Pat Reeve, Nikita Kolosov, FinnGen, Hensin Tsao, Aarno Palotie, Samuli Ripatti, Mark J. Daly, Sameer Gupta, Mykyta Artomov

## Abstract

Hypothyroidism has been associated with increased skin cancer risk, whereas greater genetic susceptibility to hypothyroidism is associated with lower risk. These opposing associations suggest that the average risk associated with hypothyroidism may conceal substantial heterogeneity among patients, and that genetic background could help identify those at highest risk. In this study, we analyzed more than 650,000 individuals across All of Us and FinnGen cohorts to determine whether a hypothyroidism polygenic risk score (H-PRS) stratifies skin cancer risk among patients with autoimmune hypothyroidism and whether integration of a skin cancer polygenic risk score (SC-PRS) further refines risk. In FinnGen, autoimmune hypothyroidism was associated with increased skin cancer hazard (hazard ratio [HR] = 1.10; 95% CI [1.06, 1.15]), whereas H-PRS was inversely associated with risk (HR=0.97 per 1-SD increase; 95% CI [0.95, 0.98]). The hypothyroidism×H-PRS interaction was also negative (HR=0.94; 95% CI [0.91, 0.98]), indicating greater hypothyroidism-associated risk at lower H-PRS. The hypothyroidism and H-PRS associations replicated in All of Us, and similar H-PRS risk gradients were observed for melanoma, basal cell carcinoma, and squamous cell carcinoma in both cohorts. Among hypothyroid patients, predicted 10-year risk increased with higher SC-PRS and lower H-PRS. Combined clinical-genetic profiles had HRs ranging from 2.32 at the 20th H-PRS/80th SC-PRS percentiles to 3.37 at the 5th/95th percentiles, exceeding published point estimates for the established dermatological risk factors. Among women aged 55, representing the more commonly affected sex at approximately the median age of diagnosis, 10-year risk of skin cancer was 14.9% (95% CI [8.4%, 20.9%]) in the highest-SC-PRS/lowest-H-PRS quartiles versus 2.8% (95% CI [1.0%, 4.7%]) in the opposite quartiles. Overall, these findings support PRS-guided risk stratification for targeted skin cancer prevention and surveillance.

## Introduction

Skin cancers constitute roughly a third of all cancer cases, and the prevalence and incidence of skin cancers have been increasing in recent decades^1^. Melanoma, squamous cell carcinoma (SCC), and basal cell carcinoma (BCC) are the three major malignancy subtypes accounting for more than 90% of all incidents of skin cancer^2^. While the leading risk factor is UV exposure^3^, other factors contribute to the development of skin cancers as well. For instance, immunosuppression is a well-established risk factor for SCC and BCC, and has also been associated with an increase in melanoma risk^4^, as illustrated by the high malignancy rates in organ transplant recipients^5^ and patients taking immunosuppressive medication^6^.

Hypothyroidism is a condition in which the thyroid gland does not produce sufficient thyroid hormones and, in iodine-sufficient populations, is most commonly autoimmune in origin^7^. It is associated with multiple skin disorders^8^, and it has been considered a risk factor for various cancers^9^. In case of skin cancer association, the existing evidence is conflicting. On the one hand, clinical studies suggest increased incidence of skin cancer to be associated with hypothyroidism^10–12^, with functional studies elaborating on the mechanism by linking thyroid hormones to tumor dynamics^8,12,13^. On the other hand, large scale genetic analyses observe inverse correlation between genetic predisposition to hypothyroidism and skin cancers^14,15^.

This discordance suggests that clinical hypothyroidism and inherited liability to hypothyroidism may capture distinct biological processes rather than alternative measurements of the same exposure. Clinical hypothyroidism reflects a realized disease state shaped by thyroid hormone deficiency, autoimmunity, treatment, comorbidities, and environmental context. In contrast, inherited susceptibility to hypothyroidism may reflect heterogeneous genetic pathways possibly independent of clinical thyroid disease. Thus, the discordance between clinical and genetic associations may reflect biological heterogeneity rather than true inconsistency, suggesting that clinical hypothyroidism and inherited liability provide complementary information for skin cancer risk stratification.

Using clinical and genetic data from more than 650,000 participants in FinnGen and All of Us, we jointly evaluated skin cancer risk as a function of diagnosed autoimmune hypothyroidism (AIHT), genetic predisposition to hypothyroidism, and their interaction. Among individuals with AIHT, lower H-PRS was associated with higher skin cancer risk in both cohorts and across skin cancer types, indicating substantial genetic heterogeneity within the diagnosed population. We then combined H-PRS with a skin cancer polygenic risk score (SC-PRS) to further stratify risk. Predicted 10-year risk increased with higher SC-PRS and lower H-PRS, and empirical analysis showed a more than fivefold difference in 10-year skin cancer risk between AIHT patients in opposing H-PRS and SC-PRS quartiles. Combined clinical-genetic profiles also showed associations comparable to or exceeding published estimates for established risk factors, including family history, UV exposure, tanning, and sunburn history. Together, these findings show that genetic information can identify substantial differences in skin cancer risk among patients with AIHT.

## Results

### Description of cohorts used for analysis

The FinnGen cohort contained 500,186 individuals of Finnish ancestry, with median age of 60.5 years, that had genetic, demographic, and longitudinal clinical data available. It included 54,083 cases of autoimmune hypothyroidism (AIHT) satisfying all of the inclusion criteria (**Methods. Exposures**). An additional 10,024 individuals had a hypothyroidism diagnosis but did not pass all of the inclusion criteria – these individuals were used for sensitivity analysis to compare how the definition of hypothyroidism affects our findings. The FinnGen cohort also included 4,563, 4,676, and 26,646 cases of melanoma, squamous cell carcinoma (SCC) of skin, and basal cell carcinoma (BCC) of skin, respectively (**Methods. Outcomes**).

The All of Us cohort comprised genetic and longitudinal clinical information for 155,186 individuals of European ancestry (median age 63.6 years). Autoimmune hypothyroidism was present in 24,256 individuals after excluding iodine-related, subclinical, post-infectious, post-operative, and medication-induced hypothyroidism cases (**Methods. Exposures**). There were 12,793 skin cancer cases, including 2,504 melanoma cases, 4,383 SCC cases, and 8,784 BCC cases (**Methods. Outcomes**).

### Brief overview of statistical analysis design

To consider the temporal information about the onset of both AIHT and skin cancer, we applied a sex-stratified Cox proportional hazards model with time-varying exposure. The primary exposures were AIHT, the hypothyroidism polygenic risk score (H-PRS), and their interaction. Models were stratified by sex and adjusted for genetic predisposition to skin cancer using the best-performing skin cancer PRS in FinnGen^16^. To partially account for geographic latitude, which may confound associations through differences in UV exposure, we also adjusted for genetic principal components (PCs) serving as proxies for geographic location^17^. Genetic PCs and PRSs were time-invariant, whereas AIHT status was modeled as time-varying and switched from 0 to 1 at the time of diagnosis (**Methods. Covariates**). Hazard ratios (HRs) greater than 1 indicate an increased risk associated with an exposure, while HRs less than 1 indicate a decreased risk. The fitted models were subsequently used to generate predicted risk curves for individuals with specified covariates (**Methods. Association test using Cox model**).

### Genetic susceptibility to hypothyroidism stratifies skin cancer risk and reflects autoimmune background

Using survival analysis, we evaluated the effects on skin cancer for three exposures in FinnGen: AIHT, H-PRS, and their interaction. At mean H-PRS, AIHT was a statistically significant risk factor for skin cancer (HR = 1.101, 95% CI [1.056, 1.148]) while H-PRS was inversely associated with skin cancer risk (HR = 0.965, 95% CI [0.953, 0.976]), with the latter effect consistently present in both AIHT cases and controls. Because this association was estimated after adjustment for AIHT, it suggests that H-PRS captures skin cancer-related biological processes that are not fully explained by diagnosed hypothyroidism. The interaction term was significantly negative (HR = 0.942, 95% CI [0.906, 0.979]), indicating that the hazard ratio associated with AIHT increasesas H-PRS decreases (**Table 1**). The sensitivity analysis (**Methods. Sensitivity Analysis**) showed that additional adjustment for BMI and smoking, restriction to a post-enrollment (prospective) design, relaxation of the hypothyroidism inclusion criteria, and addition of in situ melanoma and carcinoma to outcomes yielded effect estimates consistent with the primary analysis (**Supplementary Table 1**). The AIHT and H-PRS effects replicated in All of Us, while the interaction was directionally consistent and did not differ significantly between cohorts (**Supplementary Table 2**).

**Table 1.** Estimated effect of AIHT, H-PRS, and their interaction on the hazard of developing skin cancer in FinnGen. The analysis was performed by a stratified cox model with time-varying covariates, and p-values were calculated from Wald’s tests. The results for the main exposures are highlighted in bold.

| Variables | Hazard Ratio | 95% CI | P-value |
| --- | --- | --- | --- |
| **AIHT** | **1.101** | **[1.056, 1.148]** | **$7.2 \times 10^{-6}$** |
| **H-PRS** | **0.965** | **[0.953, 0.976]** | **$3.3 \times 10^{-9}$** |
| **H-PRS x AIHT** | **0.942** | **[0.906, 0.979]** | **$2.2 \times 10^{-3}$** |
| SC-PRS | 1.554 | [1.537, 1.572] | $< 2 \times 10^{-16}$ |
| PC1 | 0.981 | [0.970, 0.990] | $9.8 \times 10^{-5}$ |
| PC2 | 0.941 | [0.931, 0.950] | $< 2 \times 10^{-16}$ |

Together, the H-PRS main effect and its interaction with AIHT indicated substantial genetic heterogeneity in skin cancer risk among hypothyroid individuals. Among individuals with AIHT in FinnGen, those with low H-PRS values had estimated HR for skin cancer above 1.30, while those with high H-PRS had lower risk of skin cancer than the general population (**Figure 1 A**). A similar H-PRS gradient was observed in All of Us, where estimated skin cancer HRs ranged from 1.28 to 1.96 across the H-PRS distribution. (**Figure 1 B**). Moreover, similar trends were observed across the three major skin cancer types – basal cell carcinoma, squamous cell carcinoma, and melanoma – in both FinnGen and All of Us datasets (**Supplementary Figure 1**).

**Figure 1.**
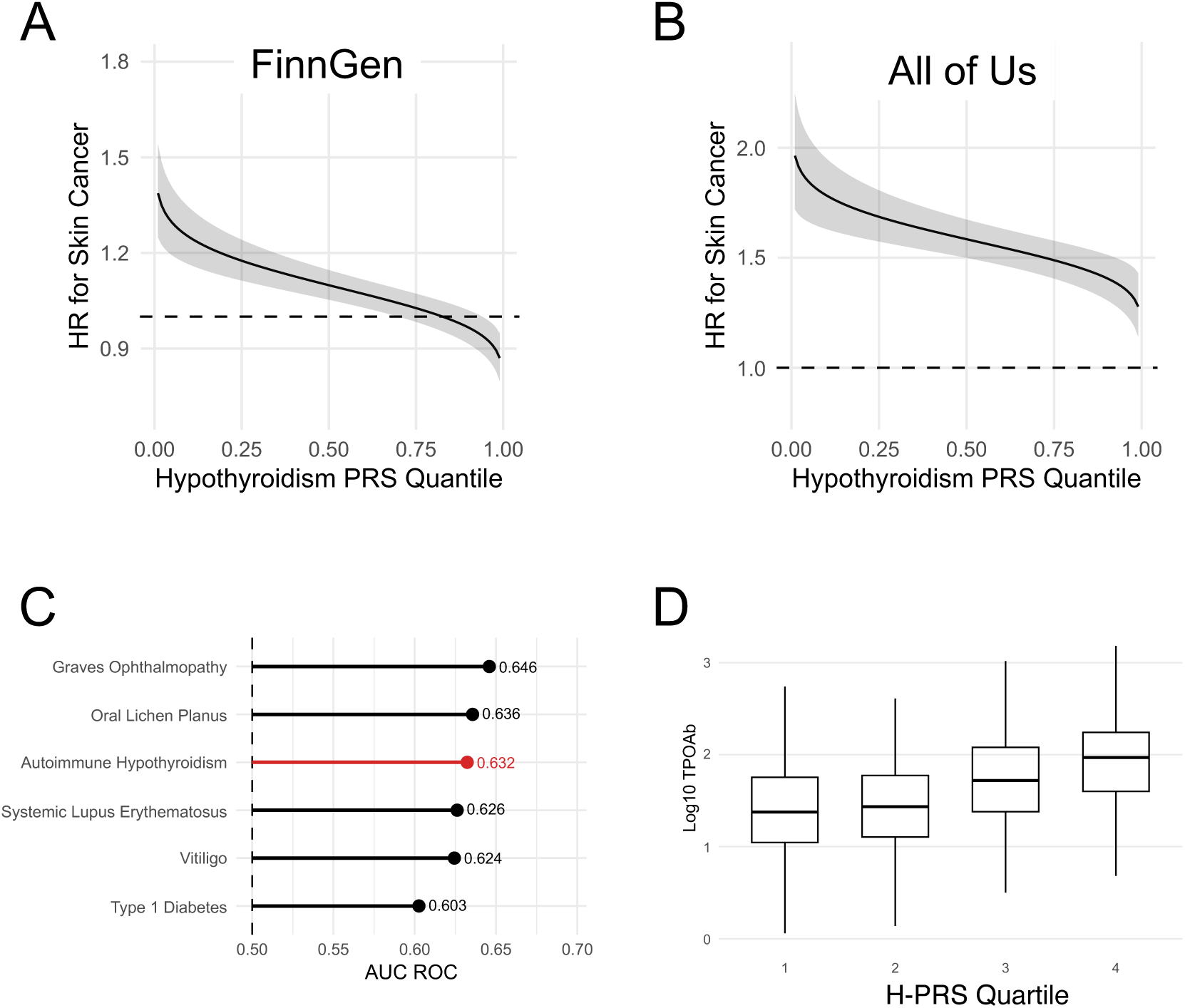
Associations of H-PRS with skin cancer risk and autoimmune phenotypes. **(A-B)** Estimated HR for skin cancer among AIHT patients in FinnGen (A) and All of Us (B) across the H-PRS distribution (shaded band: 95% CI). **(C)** Phenotypes most strongly predicted by H-PRS in FinnGen, summarized by AUROC. **(D)** Association between H-PRS quartile and thyroid peroxidase antibody (TPO Ab) levels among AIHT patients.

A major challenge in interpreting these results biologically is that H-PRS is a “black-box” biomarker, calculated as a weighted sum across nearly one million variants whose individual effects are not readily interpretable. To better characterize the biological signal represented by H-PRS, we performed a phenome-wide scan of its associations across FinnGen disease endpoints (**Methods. Analysis of correlates of polygenic risk scores**). H-PRS was associated with both broad autoimmune susceptibility and thyroid-specific autoimmunity. Specifically, H-PRS was strongly associated with multiple autoimmune diseases affecting different tissues and organs, including Graves ophthalmopathy (AUROC = 0.65, P = 1.8 × 10^−38^), oral lichen planus (AUROC = 0.64, P = 1.2 × 10^−19^), systemic lupus erythematosus (AUROC = 0.63, P = 8.0 × 10^−10^), vitiligo (AUROC = 0.62, P = 2.1 × 10^−14^), and type 1 diabetes (AUROC = 0.60, P = 1.8 × 10^−104^), among others (**Figure 1C**). Among AIHT patients, H-PRS also predicted thyroid peroxidase antibody (TPO Ab) levels: in a mixed linear model, higher H-PRS was associated with higher TPO Ab burden (N=2,386, β = 0.29, P = 9.2 × 10^−26^; **Figure 1D**).

The inverse association between H-PRS and skin cancer was consistent with a broader relationship between autoimmune genetic susceptibility and cancer risk. H-PRS was inversely associated with breast cancer (OR = 0.96, P = 4.2 × 10^−6^), prostate cancer (OR = 0.97, P = 3.6 × 10^−5^), and colorectal cancer (OR = 0.96, P = 3.3 × 10^−5^), while PRS for other autoimmune diseases, including type 1 diabetes (PGS001817; OR = 0.974, P = 5.0 × 10^−5^) and rheumatoid arthritis (PGS002769; OR = 0.963, P = 7.2 × 10^−9^), were also inversely associated with skin cancer. Because autoimmune PRS can be strongly driven by the major histocompatibility complex (MHC/HLA) region, we repeated the analysis after excluding variants from this locus. The associations between PRS and outcomes in this analysis remained significant, indicating that the inverse relationship was not explained solely by MHC/HLA variation (**Supplementary Table 3**). Together, these findings support H-PRS as a marker of autoimmune genetic susceptibility and provide a potential biological context for its inverse association with skin cancer risk.

### Translating genetic heterogeneity into skin cancer risk stratification

The preceding analyses showed that H-PRS distinguishes skin cancer risk among patients with AIHT. We next examined how this information could be used to identify high-risk individuals within this clinical group. When genotypic data are available, risk assessment can also incorporate inherited susceptibility to skin cancer itself. We therefore evaluated H-PRS jointly with a skin cancer polygenic risk score (SC-PRS), previously included as an adjustment covariate, to assess their combined potential for risk stratification.

Using All of Us, we translated this combined genetic information into age- and sex-specific estimates of 10-year skin cancer risk. We selected 60-year-old males and 55-year-old females with AIHT as illustrative profiles, roughly corresponding to the median ages of AIHT onset in males and females, respectively^18^. For each profile, we calculated risk across combinations of H-PRS and SC-PRS at their 20th, 40th, 60th, and 80th percentiles (**Figure 2A-B**). Predicted risk ranged from 7.4% to 15.2% in males and from 3.5% to 7.3% in females, with the highest estimates observed for low H-PRS combined with high SC-PRS. At each SC-PRS value, lower H-PRS was associated with higher predicted risk, providing additional differentiation beyond genetic susceptibility to skin cancer alone (**Methods. Sex- and age-specific 10-year absolute risk calculation**).

**Figure 2.**
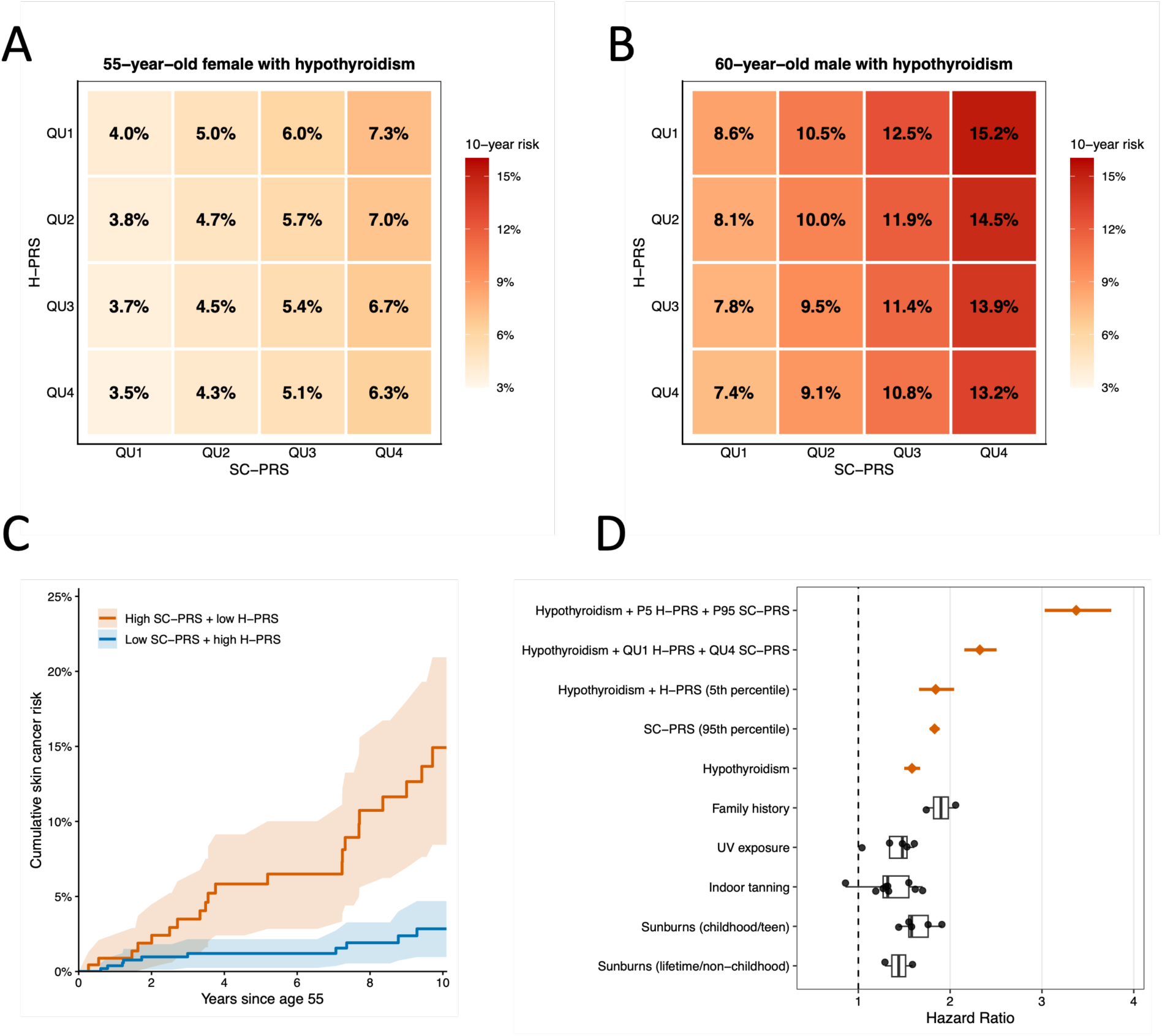
Genetic stratification of skin cancer risk among individuals with hypothyroidism. **(A-B)** Predicted 10-year risks for hypothyroid females aged 55 (A) and males aged 60 (B), across hypothyroidism (H-PRS) and skin cancer (SC-PRS) polygenic scores. QU1-QU4 denote the 20th, 40th, 60th, and 80th percentiles. **(C)** Kaplan-Meier-estimated cumulative risk curves for skin-cancer-free hypothyroid females at age 55, comparing highest SC-PRS/lowest H-PRS quartiles (orange) with opposite quartiles (blue); shading indicates 95% confidence intervals. **(D)** Study hazard ratios and 95% confidence intervals (orange) versus published risk-factor estimates (black points, summarized by boxplots). Study estimates use a non-hypothyroid reference with both PRSs set to zero. P5/P95 denote fifth/95th percentiles, QU1/QU4 denote 20th/80th percentiles.

We evaluated polygenic stratification empirically among women aged 55 with AIHT and no prior skin cancer, a group chosen because AIHT is more common in women and 55 years approximates the median age at diagnosis^18^. The Kaplan-Meier-estimated 10-year risk was 14.9% (95% CI [8.4%, 20.9%]) in the high-SC-PRS/low-H-PRS group, compared with 2.8% (95% CI [1.0%, 4.7%]) in the low-SC-PRS/high-H-PRS group (**Figure 2C, Methods. Sex- and age-specific 10-year absolute risk calculation**). This corresponded to a 5.2-fold difference in estimated risk and an absolute difference of 12.1 percentage points, illustrating substantial risk heterogeneity among patients sharing the same diagnosis, sex, and age.

To contextualize the magnitude of these associations, we compared hazard ratios associated with clinical and genetic profiles from our analysis with published estimates for established skin cancer risk factors, including family history, UV exposure, indoor tanning, and sunburn history (**Methods. Derivation of hazard ratio scale**). We assembled estimates from seven large-scale analyses of skin cancer risk factors^19–25^, including a meta-analysis of 60 studies for melanoma^19^, a meta-analysis of 38 studies for BCC^21^, and several studies evaluating risk factors for SCC^22–24^ (**Supplementary Table 4**). Additionally, we calculated effect size for the best-performing skin cancer PRS from PGS Browser^16^. For comparability across studies, published OR, RR, and HR estimates were placed on an approximate common scale for contextual comparison (**Methods. Effect-size harmonization**). Relative to a non-hypothyroid profile with reference PRS values, AIHT at reference PRS values was associated with an HR of 1.58 (95% CI [1.50, 1.67]). The corresponding HR for AIHT with H-PRS at the fifth percentile was 1.84 (95% CI [1.66, 2.04]), similar to the association for SC-PRS at the 95th percentile without hypothyroidism (HR = 1.83, 95% CI [1.78, 1.88]) and within the range of published point estimates for family history of melanoma (1.74 - 2.06). Combining AIHT with both low H-PRS and high SC-PRS identified profiles with larger relative associations. The HR was 2.32 (95% CI [2.15, 2.51]) at the 20th percentile of H-PRS and 80th percentile of SC-PRS, increasing to 3.37 (95% CI [3.03, 3.76]) at the fifth and 95th percentiles, respectively. Both estimates exceeded the published point estimates for established risk factors included in this comparison (**Figure 2D**). Together, the model-based profiles and observed risk differences illustrate how genetic information about both AIHT and skin cancer can distinguish risk among patients whose susceptibility is not fully characterized by a hypothyroidism diagnosis alone.

## Discussion

The relationship between hypothyroidism and skin cancer risk has been examined in prior observational and genetic studies, yet the overall evidence remains inconsistent. Clinical observational studies have reported modestly increased skin cancer risk among individuals with hypothyroidism, whereas large-scale genetic analyses have found a modestly protective association between genetic liability to hypothyroidism and skin cancer. Our findings provide a potential explanation for this discordance: among individuals with hypothyroidism, skin cancer risk varied according to inherited susceptibility to hypothyroidism, with lower H-PRS associated with higher risk in both FinnGen and All of Us. Thus, genetic liability to hypothyroidism provides information about skin cancer risk that is not captured by diagnosis alone.

This distinction is important because a clinical diagnosis and inherited susceptibility represent different aspects of disease. Clinical hypothyroidism reflects a realized disease state shaped by genetic, autoimmune, hormonal, treatment-related, and environmental factors, whereas H-PRS captures inherited liability to developing the condition. Patients who develop hypothyroidism despite relatively low inherited liability may therefore differ from those with high genetic susceptibility in other determinants of disease. The inverse H-PRS association among hypothyroid patients is consistent with such heterogeneity, although the observational design does not establish the underlying causal mechanism.

More broadly, these findings illustrate the context dependence of genetic effects. Genetic risk is often summarized using additive models in which the contribution of inherited susceptibility is assumed to be independent of clinical or environmental background. However, interactions can arise when the effect of genetic liability differs across disease states or other exposures^26,27^. In our analysis, the significant AIHT×H-PRS interaction indicates departure from multiplicative effects on the hazard scale: the association of AIHT with skin cancer was stronger at lower H-PRS and attenuated as inherited susceptibility to hypothyroidism increased. This pattern is conceptually related to gene-by-environment and other non-additive effects, in which the phenotypic consequence of genetic variation depends on the context in which it occurs^28,29^. Such context dependence may be particularly relevant for risk prediction, because the marginal effect of a PRS estimated across an entire population may not accurately represent its effect within clinically defined subgroups.

We further showed that this disease-context genetic information can be combined with inherited susceptibility to skin cancer itself. Predicted 10-year risk increased with higher SC-PRS and lower H-PRS, with H-PRS providing additional differentiation at each level of SC-PRS. Importantly, this pattern was also reflected in observed outcomes: among females with AIHT and no prior skin cancer at age 55, representing the more commonly affected sex at approximately the median age of hypothyroidism diagnosis, estimated 10-year risk was 14.9% in the highest-SC-PRS/lowest-H-PRS quartiles and 2.8% in the opposite quartiles. These findings demonstrate substantial risk heterogeneity within a clinically defined population. Associations for selected clinical-genetic profiles were also comparable to or larger than published estimates for established dermatological risk factors, although these cross-study comparisons involve different populations, outcomes, exposure definitions and should therefore be interpreted as contextual rather than direct comparisons of predictive performance.

The biological correlates of H-PRS provide a potential context for these associations. H-PRS was associated with multiple autoimmune diseases and with thyroid peroxidase antibody levels among individuals with hypothyroidism, suggesting that it may capture both broad autoimmune susceptibility and thyroid-specific autoimmunity. H-PRS was also inversely associated with several other cancers, while PRSs for type 1 diabetes and rheumatoid arthritis were inversely associated with skin cancer. Together with prior variant-level analyses implicating shared immune-related genetic components^15^, these findings are consistent with broader immune mechanisms linking autoimmune susceptibility and cancer risk^30^.

Several limitations should be considered. First, FinnGen consisted of individuals of Finnish ancestry and the All of Us replication analysis was restricted to participants of European ancestry, limiting generalizability to other populations. Second, detailed measures of conventional dermatologic risk factors, including cumulative UV exposure and skin phototype, were not comprehensively available, leaving the possibility of residual confounding. Third, the observational design can show predictive utility of genetic and clinical biomarkers but cannot determine whether the associations are causal. Finally, the absolute-risk analyses and PRS thresholds were used to illustrate risk stratification rather than to establish clinically validated decision thresholds.

Overall, these results show that patients sharing a diagnosis of autoimmune hypothyroidism can have substantially different future skin cancer risks depending on their genetic background. Integrating genetic susceptibility to both hypothyroidism and skin cancer provides additional risk differentiation beyond either source of information alone, potentially improving targeted skin cancer prevention or surveillance.

## Methods

### Study Participants

FinnGen (https://www.finngen.fi/en) is a public-private research project launched in 2017 with the goal of providing novel medically and therapeutically relevant insight into human diseases. FinnGen is a pre-competitive partnership of Finnish biobanks and their background organizations (universities and university hospitals) and international pharmaceutical industry partners and Finnish biobank cooperative (FINBB). All FinnGen partners are listed here: https://www.finngen.fi/en/partners. Its 13^th^ release comprises detailed genetic and longitudinal data for 519,972 Finns. After including only individuals of Finnish ancestry that had information on the necessary covariates available, we were left with a cohort of 500,186 individuals that was used for the final analysis.

For validation, we utilized the All of Us data, which was accessed through the Researcher Workbench. We only considered individuals with available disease and genotypic data, restricting the analysis to individuals of European inferred ancestry to match the original FinnGen population. This resulted in a sample of size 155,186 that was used for validation.

### Exposures

Two exposures were considered: AIHT and genetic predisposition to hypothyroidism, represented as a polygenic risk score. Diagnosis of hypothyroidism in FinnGen was corroborated by at least 3 purchases of levothyroxine. Additionally, individuals with prior thyrotoxicosis, thyroidectomy, iodine deficiency, and individuals with pituitary diseases were excluded to ensure a more homogeneous phenotype. A comprehensive description of the AIHT phenotype curation can be found in Reeve et al., 2026. Overall, 54,083 individuals had a hypothyroidism diagnosis satisfying the inclusion/exclusion criteria. Genetic predisposition was measured using a polygenic risk score from PGS Catalog that performed the best in terms of ROC AUC in FinnGen. The weights for this PRS can be found here - https://www.pgscatalog.org/score/PGS002702/. In FinnGen, individual polygenic risk scores were calculated using plink2^15,31^.

Because the registry and medication data available in FinnGen could not be reproduced exactly in All of Us, we constructed an analogous AIHT phenotype using available data to approximate the FinnGen definition. Specifically, the AIHT group included 24,256 individuals with diagnosed hypothyroidism that was not iodine-related, subclinical, post-infectious, post-operative, or medication-induced. Genetic predisposition was quantified using the same hypothyroidism polygenic risk score (H-PRS), calculated within the All of Us environment using the specialized AoUPRS package. All necessary covariates, including principal components, sex, and a skin cancer PRS, were obtained from the available All of Us datasets.

### Outcomes

The combined skin cancer endpoint included 31,988 cases with ICD-10 codes C43 (Malignant melanoma of skin) and C44 (Other malignant neoplasms of skin). Additionally, three major types of skin cancers – melanoma (ICD-10 code C43), squamous cell carcinoma (ICD-10 codes C44.02, C44.12, C44.22, C44.32, C44.42, C44.52, C44.62, C44.72, C44.82, C44.92), basal cell carcinoma (ICD-10 codes C44.01, C44.11, C44.21, C44.31, C44.41, C44.51, C44.61, C44.71, C44.81, C44.91) – were analyzed separately and had 4,563, 4,676, and 26,646 cases, respectively.

In All of Us, skin cancer cases (melanoma, BCC, SCC) were identified from diagnosed conditions within the health records. There were 12,793 skin cancer cases, including 2,504 melanoma cases, 4,383 SCC cases, and 8,784 BCC cases.

### Covariates

We used the first two genetic principal components, sex, and polygenic risk score for skin cancer as covariates. Principal components were computed from the genotypic FinnGen data using PLINK 2.0 and were shown to correlate with the geographic location in FinnGen^17^. We used the skin cancer PRS from PGS catalogue that had the best ROC AUC performance in FinnGen, and the weights for this PRS can be found here - https://www.pgscatalog.org/score/PGS002014/. PRS was used to adjust for potential discrepancy in genetic susceptibility to skin cancer between hypothyroid cases and controls. For All of Us dataset, PRS was calculated using the AoUPRS package^32^.

### Association test using Cox model

Cox proportional-hazards model with time-varying exposure was used for the main analysis. It is implemented with the *coxph* function in the *survival* package. To allow for the time-varying covariates, we augmented our data using the *tmerge* function from the same *survival* package. Overall, genetic-based covariates like principal components or skin cancer PRS were constant throughout life, while the hypothyroidism status changed from 0 to 1 at the time of diagnosis. Additionally, the analysis was stratified by sex to allow different baseline hazards for males and females. The final model can be described as Skin cancer ∼ AIHT + H-PRS + AIHT ×H-PRS + Skin cancer PRS + PC1 + PC2 + strata(Sex) + Non-autoimmune hypothyroidism. Individuals with non-autoimmune hypothyroidism were retained in the analysis but modeled with a separate time-varying indicator. This allowed their follow-up and skin cancer events to contribute information for estimating the effects of shared covariates while preventing non-autoimmune hypothyroidism from being included in the AIHT exposure group. For a reference of survival analysis using time-varying covariates see https://stat.ethz.ch/R-manual/R-patched/library/survival/doc/timedep.pdf.

### Sensitivity Analysis

We performed four sensitivity analyses to assess robustness of the primary findings. First, to evaluate potential confounding by major lifestyle factors, we repeated the primary time-varying Cox model in the subset with available body mass index (BMI) and smoking information, additionally adjusting for BMI and smoking, and compared hazard ratios (HRs) for clinical hypothyroidism, the hypothyroidism polygenic risk score (H-PRS), and their interaction with the main analysis. Second, to assess sensitivity to our stringent autoimmune hypothyroidism definition, we expanded the exposure to include 10,024 additional hypothyroidism cases that did not meet the autoimmune criteria (e.g., levothyroxine purchase requirements and thyroid-related exclusions) into the AIHT group. Third, to align the analysis with a prospective post-enrollment design and reduce reliance on pre-enrollment outcome ascertainment, we restricted follow-up to begin at FinnGen enrollment age (BL_AGE) and counted only skin cancer events occurring after enrollment (with hypothyroidism modeled as in the primary analysis), noting that this restriction reduces the number of events and therefore precision. Fourth, we included in situ melanoma and carcinoma to skin cancer definition to see how a wider definition of the outcome affects the effect size estimates.

### Analysis of correlates of polygenic risk scores

The associations between polygenic risk scores and diseases in FinnGen were obtained from PGS Browser – a resource with pre-computed ROC AUC, odds ratio, and p-values for associations between 3,168 PRS models and 4,739 traits^16^. The results for polygenic risks calculated with variants from MHC locus excluded were also obtained from PGS Browser. The association between H-PRS and TPO Ab was tested with a linear mixed model implemented in the `lme4` R package, with a participant-level random intercept to account for repeated measurements. The outcome was the logarithm of TPO Ab concentration measured in iu/ml. This analysis was adjusted by sex, age of measurement, whether the test was taken from serum or plasma.

### Sex- and age-specific 10-year absolute risk calculation

Ten-year Cox model-based skin cancer risk was estimated in All of Us from the fitted Cox proportional-hazards model, using age as the analysis time scale and sex-specific baseline hazards. For an individual with covariate vector *x*, conditional survival from age *t* to age *t* + 10 was estimated as

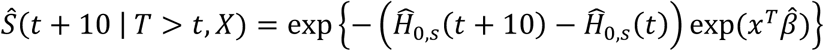

here *Ĥ*_0,*s*_ is the estimated cumulative baseline hazard for sex *s* obtained via `basehaz(centered = FALSE)` R function, and *x*^*T*^*β̂* is the linear predictor from the fitted Cox model.

The empirical 10-year skin cancer risk was estimated using the Kaplan-Meier estimator implemented with the `survfit` function in the R `survival` package. Skin-cancer-free survival *Ŝ*_*g*_(*t* + 10 | *T* > *t*) was estimated separately for each genetic group, and then the risk was estimated as *R̂*_*g*_(*t* + 10 | *T* > *t*) = 1 − *Ŝ*_*g*_(*t* + 10 | *T* > *t*). Pointwise 95% confidence intervals for 10-year risk were obtained by transforming the corresponding Kaplan-Meier confidence limits for survival.

### Derivation of hazard ratio scale

To calculate hazard ratios with confidence intervals corresponding to individuals with diagnosed hypothyroidism and particular PRS quantiles, we obtained estimated Cox model coefficients *β̂* using `coef()` function, and the covariance matrix *Σ* of this estimate using the `vcov()` function in R.

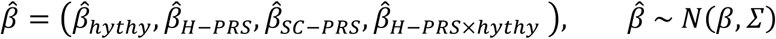

Then, we can define the vector of features for a patient with 1 corresponding to hypothyroidism diagnosis, and *Φ*^−1^(*q*_*H*−*PRS*_) and *Φ*^−1^(*q*_*SC*−*PRS*_) corresponding to the standardized PRS value for quantile *q*:

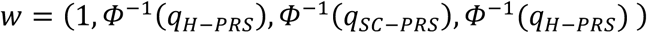

Then, the estimated log hazard ratio 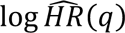 for this patient is equal to *w*^*T*^*β̂*, and has a normal distribution:

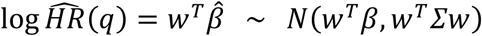

Finally, from this normal distribution, we can calculate 100(1 − *α*)% confidence intervals as:

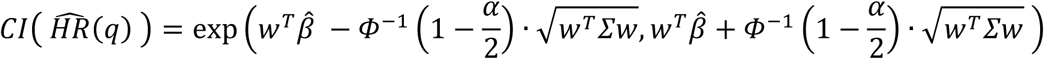

### Effect-size harmonization

Published associations for established skin cancer risk factors were reported using heterogeneous measures (odds ratios (OR), relative risks (RR), and hazard ratios (HR)). To compare them on a common scale, we mapped non-HR measures to an approximate HR scale assuming that the risk of skin cancers over the relevant time horizon is small (*p*_0_ ≪ 1). When cumulative incidence is small, OR and RR are numerically close because 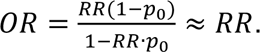 In addition, when incidence is low (thus survival is high *S* (*t*) = 1 − *p*_0_ ≈ 1) and under a proportional hazards model with proportional hazard, 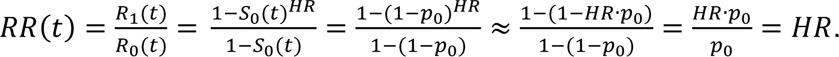 Thus, HR can be approximated as RR. As such, for diseases with low baseline risk *HR* ≈ *OR* ≈ *RR*^33^. Accordingly, for the purpose of contextualizing effect sizes, we treated OR and RR estimates from the literature as approximate HRs and compared them to HRs estimated in our Cox models. This harmonization was used for benchmarking only and does not affect inference in the primary survival analyses.

### Ethics statement

FinnGen. Individuals in FinnGen provided informed consent for biobank research, based on the Finnish Biobank Act. Alternatively, separate research cohorts, collected prior to the Finnish Biobank Act came into effect (in September 2013) and the start of FinnGen (August 2017), were collected based on study-specific consents and later transferred to the Finnish biobanks after approval by Fimea (Finnish Medicines Agency) and the National Supervisory Authority for Welfare and Health. Recruitment protocols followed the biobank protocols approved by Fimea. The Coordinating Ethics Committee of the Hospital District of Helsinki and Uusimaa (HUS) statement number for the FinnGen study is Nr HUS/990/2017. Further details on permit and biobank decision numbers are available in the FinnGen Ethics Statement in the Supplementary Information.

All of Us participants provided informed consent for participation in the Research Program. The All of Us protocol and participant-facing materials were approved by the All of Us Institutional Review Board. The present study used Controlled Tier data through the Researcher Workbench; individual projects using these data do not require review by the All of Us IRB.

## Supporting information

FinnGen author list

## Data availability

The individual-level FinnGen data used in this study were obtained under FinnGen Proposal Number F_2020_073 (https://www.finngen.fi/en). The FinnGen data are not publicly available because they contain sensitive genetic and health information protected by participant consent, ethical approval, and Finnish and European data protection regulations, including the General Data Protection Regulation (GDPR). Thus, the raw FinnGen data are protected and are not publicly available due to data privacy laws. Details on the data-access restrictions and policies are listed within the FinnGen flagship publication - Kurki et al., Nature, 2023. This study used data from the All of Us Research Program’s Controlled Tier Dataset version 8, available to authorized users at https://www.researchallofus.org/.

## Acknowledgements

We acknowledge the contribution of FinnGen and All of Us Research Program and their participants to this work.

## Author contributions

I.M., M.A. designed and conceived the study. N.K. and FinnGen contributed data and analysis pipelines. I.M. conducted the analysis and generated figures. I.M. and M.A. wrote the manuscript. M.P.R., H.T., S.G. contributed to the interpretation of results. M.A., M.J.D., S.R., and A.P. acquired funding. All authors reviewed and edited the manuscript.

## Funding

I.M., N.K., and M.A. disclose support for this work from the Aging Biology Foundation. The FinnGen project is funded by two grants from Business Finland (HUS 4685/31/2016 and UH 4386/31/2016) and by the following industry partners: AbbVie Inc., AstraZeneca UK Ltd, Biogen MA Inc., Bristol Myers Squibb (including Celgene Corporation and Celgene International II Sàrl), Genentech Inc., Merck Sharp & Dohme LLC, Pfizer Inc., GlaxoSmithKline Intellectual Property Development Ltd., Sanofi US Services Inc., Maze Therapeutics Inc., Janssen Biotech Inc., Novartis Pharma AG, and Boehringer Ingelheim International GmbH.

## Conflict of interest

Findings described in this work are a part of provisional patent US PTO #64/081,466. Mark J. Daly is a founder of Maze Therapeutics. The remaining authors declare no competing interests.

## Notes

**Conflict of interests statement is in the end of the manuscript.**

### Author Declarations

FinnGen. Individuals in FinnGen provided informed consent for biobank research, based on the Finnish Biobank Act. Alternatively, separate research cohorts, collected prior to the Finnish Biobank Act came into effect (in September 2013) and the start of FinnGen (August 2017), were collected based on study-specific consents and later transferred to the Finnish biobanks after approval by Fimea (Finnish Medicines Agency) and the National Supervisory Authority for Welfare and Health. Recruitment protocols followed the biobank protocols approved by Fimea. The Coordinating Ethics Committee of the Hospital District of Helsinki and Uusimaa (HUS) statement number for the FinnGen study is Nr HUS/990/2017. Further details on permit and biobank decision numbers are available in the FinnGen Ethics Statement in the Supplementary Information. All of Us participants provided informed consent for participation in the Research Program. The All of Us protocol and participant-facing materials were approved by the All of Us Institutional Review Board. The present study used Controlled Tier data through the Researcher Workbench; individual projects using these data do not require review by the All of Us IRB.

